# Cognition, physical function, and life purpose in the rural elderly population: a systematic review protocol

**DOI:** 10.1101/2023.09.06.23295116

**Authors:** Hércules Lázaro Morais Campos, Elisa Brosina De Leon, Ingred Merllin Batista de Souza, Anna Quialheiro, Elizabete Regina Araújo de Oliveira

## Abstract

**Introduction:** Aging in the rural setting world from the perspective of cognition, physical function, and life purpose, essential constructs for a prosperous old age, still needs to be better discussed. Thus, this systematic review protocol highlights the prevalence of cognitive decline, physical functioning, and life purpose in older adults aging in rural community settings. Methods and analysis: We will include cross-sectional studies published until April 2022 found in 8 databases: Embase, MEDLINE, LILACS, PsycINFO, Scopus, SciELO, and Web of Science. For the first selection of studies, the Ryyan software will be used, and to check the methodological quality and the risk of bias, we will use COSMIN. For the primary analysis, the titles and abstracts available in the search engine will be analyzed by the following MeSH descriptors “Physical functioning,”; “Cognition,”; “Cognitive function,”; “Life purpose,”; Personal satisfaction; Subjective well-being; “Elderly”; “Older”; “Rural aging”; “Rural population”; “Communities, rural”; “Distribution, rural spatial”; “Medium communities”; “Rural settlement”; “Small community”. If necessary, the secondary analysis will adopt a complete reading of the selected articles by two blinded reviewers and confirmed by a third person. Publication bias will be assessed using cross-sectional analytical study quality. Sensitivity analyses will be performed by retrieving one article at a time and analyzing their endpoints with the proposal to identify the manuscripts that significantly influenced the combined prevalence of the endpoints.

**Strengths and limitations of this study:** It will present worldwide epidemiological data on aging in rural settings from the perspective of cognition, physical functioning, and life purpose.

It is the first systematic review that involves the theme of life purpose worldwide in rural elderly.

It is a cross-sectional study that although it cannot describe clinical outcomes, presents sociodemographic, cognition, physical functioning, and life purpose data that can help in public health decision-making for this population.

## INTRODUCTION

Aging within the rural context takes place in a variety of ways around the world, and increasingly the literature has pointed out that people aging in this unique setting have different epidemiological and health indicators than seniors aging in the urban context (1)

There are controversies about the elderly aging in the rural context, sometimes the challenges are pointed as independence and active participation in community life, as well as attention to safety, choice, and location of housing, loneliness, social isolation, difficulty accessing services, leisure, food, transportation, agriculture itself until retirement. On the other hand, seniors who age in rural settings may have good health, and quality of life especially in the cognitive(1) aspect, and have better access to health, services, and healthier living and eating habits although they are at constant risk of frailty (2)

Well-conducted cross-sectional and observational studies have relevance within aging and measure the prevalence of health outcomes as well as help understand the social determinants of health and above all describe the characteristics of a given population (3)

There is still no synthesis of what aging in the rural world context looks like from the perspective of cognition, physical function, and life purpose. Thus, this systematic review protocol was designed with the following question: (1) what is the prevalence of cognition, physical functioning, and life purpose in older adults aging in rural community settings?

## METHODS AND ANALYSIS

This systematic review protocol followed the PICO strategy and was registered with the International prospective register of systematic reviews (PROSPERO) under number 2022 CRD42022311053.

### Eligibility Criteria

#### Databases and Search Strategy

The search will include cross-sectional studies until April/2022 in journals indexed in the following health databases: Embase, Medical Literature Analysis and Retrieval System Online (MEDLINE), Latin American and Caribbean Literature in Health Sciences (LILACS), PsycINFO, SciVerse Scopus (Scopus), SciELO (Scientific Electronic Library Online) and Web of Science. Published until December 2021, studies that epidemiologically evaluated the outcomes of cognition, functionality, and life purpose within rural aging and that were published in Portuguese, English or Spanish will be accepted. The protocol will follow the Preferred Reporting Items for Systematic Reviews (PRISMA) guidelines (4)

The search methods will use 3-step Boolean operators with the search strategies following the uniqueness of each database. The search strategy will use: “rural aging”, “elderly”, “old”, “elderly”, “physical functioning”, “cognition”, and “life purpose” according to Boolean operators. Two independent HLMC and EBDL reviewers will search the studies, and if necessary, a third reviewer will be included to decide on tie-breaking criteria.

#### Study selection and data extraction

After evaluation of the titles and abstracts retrieved from the searches, potential full texts will be assessed for eligibility by two independent reviewers. Authors of potential full texts will be contacted for questions about the eligibility criteria and studies will be excluded when there is no response. Studies meeting the eligibility criteria will be included. For studies with the same sample, only those with the most representative sample of that population will be considered.

Data relevant to the theme in question will be extracted from the selected articles. The selection of articles will contemplate the following eligibility criteria: (a) inclusion criteria: studies that presented a prevalence of functioning conditions, cognitive decline, and life purpose in rural elderly; and (b) exclusion criteria: systematic review studies; methodological studies; instrument validation studies; and qualitative studies. The following information will be recorded: year of publication of the study, authors, characteristics of the institutions in which the study was conducted, sampling strategy, type of sample, prevalence of outcomes, and their associated factors. The information will be organized and presented in tables.

#### Study Evaluation

To assess the methodological quality of each study, i.e., to assess the risk of bias in a study’s results, the corresponding COSMIN Risk of Bias box must be filled in. To determine the overall quality of a study, the lowest rating of any standard in the box is taken (i.e. the “worst score counts” principle). For example, if for a reliability study, one item in a box is rated as ‘inadequate’, the overall methodological quality of that reliability study is rated as ‘inadequate’ which will generate a quality score at the end of the systematic review (5).

#### Strategy for Data Overview

The synthesis of outcome study data will be organized and presented according to the recommendations of the Meta-analysis of Observational Studies in Epidemiology (MOOSE) statement (6). When two or more articles are reported as results from the outcome database, only the most comprehensive one (with more details on sampling strategy and response rate) will be included in the meta-analysis. Results will be presented in Forest Plots, showing 95% confidence intervals and p-values. Heterogeneity will be assessed using the I^2^ statistic, which will be considered high when I^2^ is equal to or greater than 75%. Statistical procedures will be performed in STATA 14.0, with the significance level set at 5% for two-tailed tests (6).

#### Patient and Public Involvement

There is no direct patient or public involvement in this study.

## ETHICS AND DISSEMINATION

Ethical approval is not required for this study since no personal or private information of the individuals will be involved. It is intended to submit this study to a peer-reviewed academic journal.

Cross-sectional studies are more economically feasible but easy to conduct and can establish preliminary evidence so that further advanced studies can be conducted. Prevalence and incidence are established and can generate important hypotheses, they can be analytical or descriptive, and depending on whether the outcome variable is assessed possible associations with exposures or risk factors can be established (3).

It seems that the functional physical capacity of elderly people aging in rural settings needs to be better understood, elderly women with good income are more likely to be sedentary as well as elderly men (7).

Studies have demonstrated the factors associated with frailty in rural elderly such as age, gender, health status variables and that included self-perception of health and number of chronic conditions, functional covariates variables such as disability in basic activities of daily living (ABVD), disability in instrumental ADLs, length of stay in the chair and psychosocial problems, it seems that depressive symptoms and cognitive impairment, greater comorbidity and disability was found in this population (8).

As for cognition, there is divergence in the literature related to the place of birth and where the rural elderly live. Sometimes the rural environment is pointed out as positive for aging (2) and at other times not (9) and (10).

The Purpose of Life in rural elderly is still little investigated and very little is known about the impact of this condition on rural aging until this moment the literature points out that the elderly with a purpose of life are more resilient, have less risk for developing dementia, feel less pain and are happier and more functional (11). There are many questions to be answered about the way of aging in the rural context and some gaps still need to be filled and understood to understand rural elder aging. After all, is rural aging positive or negative on the cognition, physical function, and life purpose of the elderly population?

## Data Availability

N/A.

## Contributions

HLMC-Writing the review protocol, reviewing the search strategies, recording the review, writing the manuscript, and final revision of the manuscript.

EBDL-Writing of the review protocol, review of the search strategies, and final revision of the manuscript.

IMBS-Writing of the review protocol, review of the search strategies and final revision of the manuscript.

AQ-Writing of the review protocol, review of the search strategies and final review of the manuscript.

ERAO-Writing of the review protocol, review of the search strategies and final review of the manuscript.

